# DIA-PINN. A physics-informed machine learning method to estimate global intrinsic diastolic chamber properties of the left ventricle from pressure-volume data

**DOI:** 10.64898/2026.03.02.26347245

**Authors:** Javier Fernández-Topham, Manuel Guerrero-Hurtado, Juan Carlos del Álamo, Javier Bermejo, Pablo Martinez-Legazpi

**Author notes:** Corresponding Author: Pablo Martinez-Legazpi, Department of Mathematical Physics and Fluids, Universidad Nacional de Educación a Distancia, Madrid, Spain.

## Abstract

**Background:** Pressure–volume (PV) loop analysis remains the gold standard for assessing the intrinsic global diastolic properties of the left ventricle (LV). Traditional fitting techniques rely on local, phase-constrained fittings and are limited due to their sensitivity to noise, landmark selection, violation of assumptions, and non-convergence.

**Objective:** To develop and validate DIA-PINN, a physics-informed neural network (PINN) framework capable of calculating intrinsic diastolic properties of the LV from measured instantaneous PV data, combining mechanistic interpretability with machine learning flexibility.

**Methods:** Instantaneous LV diastolic pressure was modeled as the sum of 1) time-dependent relaxation-related pressure and 2) volume-dependent recoil and stiffness-related pressures. DIA-PINN was trained using time, LV pressure and volume as inputs, enforcing data fidelity, model consistency, and physiological plausibility within the loss function. Performance was evaluated in 4,000 Monte Carlo simulations of LV PV-loops, and in clinical data from 59 patients who underwent catheterization (39 with heart failure and normal ejection fraction and 20 controls). DIA-PINN derived indices were compared to those obtained from a previously validated global optimization method (GOM).

**Results:** On the simulation data, DIA-PINN accurately recovered all constitutive indices (intraclass correlation coefficients near unity) and improved GOM performance. On the clinical data, diastolic indices derived using DIA-PINN strongly correlated with GOM estimates (R>0.90, p<0.001) but were insensitive to initialization. DIA-PINN performed best under vena cava occlusion, as varying preload improved parameter identifiability.

**Conclusions:** When applied to instantaneous pressure-volume data, a generalizable PINN framework, DIA-PINN, provides an improved method for assessing global intrinsic diastolic properties of cardiac chambers.

**New & Noteworthy:** Our work introduces DIA-PINN, a physics-informed neural network framework to process instantaneous ventricular pressure–volume data, solving a mechanistic model of diastole with machine learning techniques. Compared to current conventional or optimization-based approaches, the PINN provides the most reliable estimates of diastolic stiffness, relaxation, and elastic recoil, unsensitive to initialization. By embedding physiological constraints into network training, this approach achieves robust, interpretable, and clinically applicable quantification of gold-standard metrics of intrinsic global diastolic chamber properties.

## Introduction

Left ventricular (LV) diastolic dysfunction plays a major role in most cardiac diseases, such as heart failure, cardiomyopathies, and valvular heart disease (1, 2). Regardless of the underlying etiology, abnormal diastolic ventricular mechanics contribute to elevated filling pressures, pulmonary congestion and reduced cardiac performance (3, 4). Consequently, an accurate characterization of LV global intrinsic diastolic properties is essential to understand disease mechanisms. Both in clinical and experimental settings, a robust characterization of diastolic function is required to stratify patients, guide therapies, and validate novel diagnostic and therapeutic techniques in vivo (5). However, despite decades of research, we still lack of a widely generalizable method to comprehensively assess diastolic chamber mechanics across clinical and experimental settings (1, 2, 6).

Invasive instantaneous pressure–volume (PV) loop measurements are the reference standard for evaluating global intrinsic diastolic chamber properties in both ventricles (7, 8). This methodology enables direct access to mechanical properties across cardiac cycles and is very robust when combined with acute load manipulations (9).

Traditional methods for PV loop analysis rely on separately fitting specific segments of data, such as the isovolumic relaxation phase or the end-diastolic points over multiple beats. Under this methodology, exponential or logistic functions are used to fit the measured pressure and compute the time constant of relaxation (τ) or the slope of the end-diastolic pressure-volume relationship (EDPVR), respectively (10). This approach, however, has several limitations. First, it assumes that relaxation and volume-dependent properties are uncoupled as it neglects active relaxation during late diastole and volume-dependent pressure changes during early diastole. Secondly, traditional methods do not utilize the entire diastolic segment of the PV loop, increasing susceptibility to noise, beat-to-beat variability, and landmark detection errors. Moreover, they usually do not account for the restoring forces generated below the slack chamber volume (V□), which play a key role in early filling (11). Accordingly, these methods may fail in disease states associated with incomplete relaxation or altered load (9).

Some of these limitations were previously addressed by a global optimization method (GOM) that fits complete diastolic periods of multi-beat PV loops and utilizes physiologically informed constitutive equations (6). Using this methodology, we demonstrated that it is possible to decouple active time-dependent and volume-dependent diastolic forces (11-14). Furthermore, the GOM also enables the assessment of restoring pressures in intact hearts and has demonstrated greater reliability than the conventional method for single-beat measures of stiffness. The GOM has shown good accuracy across synthetic, animal, and clinical datasets, with significant improvements in reproducibility and uncertainty quantification (6). Furthermore, key aspects of diastolic function and dysfunction of both the left and right ventricles have been revealed using this GOM methodology (11, 12, 14). However, despite its advantages, the GOM is limited by its sensitivity to parameter initialization and bounds, which in some cases can result in convergence to spurious local minima (15).

In the present study we propose a further improvement of the GOM method for the analysis of global PV data by implementing a physics-informed neural network (PINN) framework for modeling LV DIAstolic mechanics, DIA-PINN. PINNs incorporate validated governing equations directly into neural network training as soft constraints, combining the interpretability of mechanistic models with the flexibility of machine learning□(16). PINNs have recently emerged as a powerful framework in cardiovascular modeling, primarily for problems involving spatially distributed fields or reduced-order circulatory dynamics. Existing applications have focused on flow reconstruction from cardiac imaging, inverse characterization of cardiac electrophysiology, or estimation of systolic parameters such as contractility within lumped-parameter or closed-loop circulation models (17-21). However, these approaches do not address the direct fitting of invasive pressure–volume loop data during diastole, nor the estimation of intrinsic diastolic chamber properties governed by coupled time- and volume-dependent mechanisms. In this context, the present work extends the scope of PINNs to a fundamentally different problem: the inference of global diastolic mechanical properties directly from instantaneous PV measurements, using a physiologically grounded constitutive model embedded within the learning process. By operating on raw diastolic PV trajectories and enforcing mechanistic consistency, this framework bridges physics-informed machine learning with gold-standard invasive assessment of diastolic function, complementing and advancing traditional fitting and optimization-based approaches.

## Methods

### DIA-PINN Framework

We based DIA-PINN on a feedforward neural network (FNN), using chamber pressure and volume waveforms vs time as inputs and predicted pressure during the entire diastole as a single output. The constitutive parameters (indices of diastolic functions) were treated as global trainable variables. The loss function guiding network training was designed to enforce three simultaneous objectives and encompasses several terms, ℒ = ℒ_*phys*_ + ℒ_*data*_. First, it incorporates a physics-informed term, ℒ_*phys*_, to ensure that the predicted pressure conforms to the underlying constitutive equations, described below. Second, a data fidelity term, ℒ_*data*_, aims to minimize the mismatch between the predicted and measured pressure waveforms. Finally, the loss function included a physiological plausibility component, defined as a ReLU-like constraints, which enforced consistency with established LV mechanical constraints to guarantee physiologically plausible results (**Figure 1**). This multi-component loss ensures physical plausibility and robustness even under sparse or noisy data.

**FIGURE 1.**
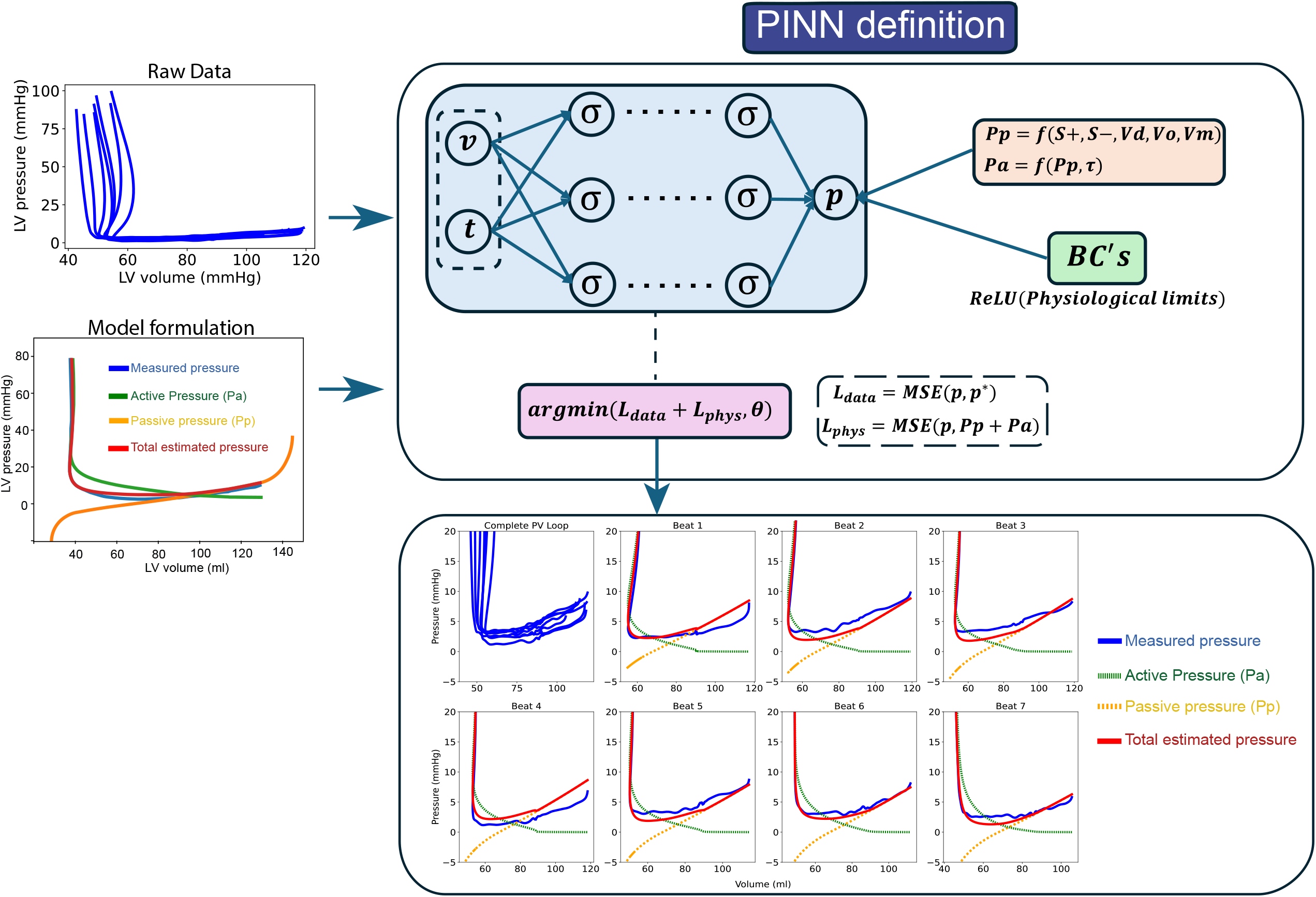
Schematic overview of the DAI-PINN framework. Instantaneous pressure–volume (PV) loop data (pressure, volume, and time) are provided as inputs to the physics-informed neural network. During training, the network predicts the diastolic pressure waveform while enforcing the underlying constitutive equations through the loss function and simultaneously fitting the measured data. The trained model yields estimates of the constitutive parameters and decomposes total diastolic pressure into its active (relaxation-related) and passive (volume-dependent) components.

The architecture and hyperparameters of the FNN were tuned using 50 synthetic cases based on real volume waveforms, as described below. In this process, we considered 1, 2, and 5 layers, and neurons and epochs varied among 1, 2, 5, 8, and 32, neurons per layer with several activation functions (Sigmoid, ReLU, ELU, and Tanh), and 10·10^3^, 50·10^3^, and 100·10^3^ training epochs. We also tested three different learning rates: 10^-2^, 10^-3^, 10^-4^ for a total of 27,000 runs. The optimal FNN configuration was selected via grid search based on the lowest loss values across all synthetic cases. Finally, we established an early stopping criterion to avoid excessive computational time and overfitting.

The optimal PINN architecture consists of only two hidden layers, each containing five neurons. Sigmoid activation functions and Adam optimizer with 50,000 epochs, with a learning rate of 0.001, also showed to be optimal parameters for the PINN. The fitting PINN algorithm was programmed in python (v 3.9) and full code can be obtained at: https://github.com/jftopham/PVLoop_PINN.

### Constitutive Model

The physical loss term in the PINN followed a previously derived constitutive model (6). Diastolic pressure was modeled as the sum of “active” and “passive” components, P_(*V*)_ = P_*a*(*t,V*)_ + P_*p*(*V*)_, following well validated constitutive equations (6, 22, 23). The active pressure component was described using an exponential decay function (23-25),

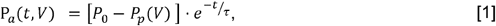

where *P*_0_ is the LV pressure at the onset of diastole, *t* is the time elapsed since onset, and T is the time constant of ventricular relaxation. Passive pressure (*P*_*p*_) was assumed to follow a piecewise logarithmic function, characterized by the coefficients of stiffness and elastic recoil, *S*_+_ and *S*_*-*_, and the equilibrium volume, *V*_0_, corresponding to *P*_*p*_ = 0,

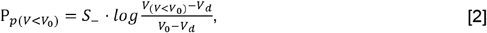

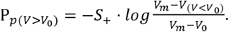

Eq [2] also depends on the constitutive parameters *V*_*d*_ and *V*_*m*_, which define the maximum and minimum left ventricular volume. Pressure continuity condition at *V*_0_ implies that

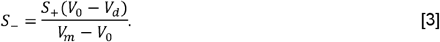

This formulations yields the diastolic pressure defined by a subspace of six constitutive parameters:

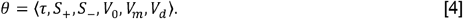

### Synthetic Data Generation and Monte Carlo Simulation

To evaluate the robustness and accuracy of the PINN, we generated synthetic datasets through Monte Carlo simulations. We used volume-time waveforms from 20 patients (see below) as templates for rescaling (14). These waveforms served as realistic input volume profiles from which pressure curves were simulated using the constitutive model described above.

In the models, all five constitutive indices governing diastolic pressure —(*τ*, *S*_+_, *V*_0_, *V*_*m*_, *V*_*d*_) — were systematically varied across physiologically plausible ranges to create a diverse synthetic population of 4,000 cases. Specifically, we varied the time constant of relaxation (T) from 35 to 75 ms, the passive stiffness constant (*S*_+_) from 5 to 82 mm Hg, the equilibrium volume from 8 to 165 mL, the dead volume asymptote, *V*_*d*_, from 2 to 65 mL, and the maximum volume asymptote (*V*_*m*_) from 60 to 300 mL. Additional parameters included a baseline end-systolic pressure (*P*_0_) between 80 and 200 mm Hg and a heart rate of 40 to 80 beats per minute. Physiological constraints imposed on the simulations included an ejection fraction between 0.25 and 0.75, *V*_0_ and *V*_*m*_ > 1.2 · *V*_*d*_ and *V*_*m*_ > 1.2 · EDV, while V_*d*_ was constrained to < 1.2 · ESV.

To synthetically reproduce a vena cava occlusion (VCO), preload was progressively reduced over five consecutive cardiac cycles while keeping the cardiac timing unchanged. Stroke volume and end-systolic volume were decreased by 1.5% per beat, and the baseline pressure parameter, *P*_0_, was reduced by 5% per beat. This procedure generated a family of PV loops with decreasing preload, consistent with the physiological response to VCO (see **Supplemental Figure SF1**).

Each diastolic pressure–volume waveform was discretized into 150 uniformly sampled collocation points for PINN training. To assess robustness, both random and Gaussian noise were added to the pressure signals across a range of amplitudes. This strategy tested the ability of the PINN to recover the underlying constitutive parameters under noisy, experimentally realistic conditions.

### DIA-PINN robustness and sensitivity analyses

The sensitivity of DIA-PINN to the nature of the input data was evaluated by comparing its performance when trained on full vena cava occlusion sequences versus isolated single-beat recordings. This analysis quantified model accuracy in the absence of preload manipulation, a common scenario in experimental and clinical acquisitions. To assess robustness to parameter initialization, a sensitivity analysis was performed using a subset of 750 synthetic PV datasets with vena cava occlusion, each trained with five independent random initializations of the parameter vector (3,750 runs in total). This procedure enabled a quantitative evaluation of training stability and the model’s ability to avoid convergence to local minima. Finally, the performance of DIA-PINN was systematically compared with that of the global optimization method under two constraint settings: physiological bounds and extended bounds obtained by doubling the parameter ranges, thereby testing robustness to broader search spaces and susceptibility to non-physiological solutions.

### Validation with Clinical Data

We evaluated the clinical performance of the PINN method using invasive PV data obtained from a well-characterized patient cohort. It comprises a total of 59 patients in sinus rhythm, referred for coronary angiography to the Department of Cardiology and Pneumology at Charité Campus Benjamin Franklin (Berlin, Germany). This patient population has been described in detail in the past (14, 26, 27). The clinical cohort included two groups. The first group consisted of 39 patients with a history of heart failure with preserved ejection fraction (HFpEF; ejection fraction > 50%), and echocardiographic evidence of diastolic dysfunction. The second group included 20 control patients with atypical chest pain or intermittent arrhythmia and normal atrial peptide values and no evidence of systolic or diastolic dysfunction by Doppler-echocardiography.

All procedures were conducted in accordance with the Declaration of Helsinki, and the research protocol was approved by the institutional ethics committee of Charité University Medicine Berlin (protocol number 225-07). Written informed consent was obtained from all participants prior to inclusion. High-fidelity PV loop recordings were acquired using 7F pigtail pressure–conductance catheters via a dual-field acquisition system (CDLeycom CFL-512). Left ventricular volumes were calibrated using the thermodilution and hypertonic saline technique. Complete hemodynamic datasets were collected during transient preload reduction via inferior vena cava occlusion. The PINN-derived parameter estimates were again compared with those obtained using the GOM (6, 14) in this dataset.

### Statistical Analyses

Multiple metrics were computed to address the accuracy, consistency, and robustness of the PINN framework. To quantify the accuracy of the PINN to recover the simulation parameters, we generated Bland–Altman plots for each of the five constitutive parameters. Results of the PINN method were compared to the GOM estimates. Results are reported in terms of bias ± standard error, as well as the intraclass correlation coefficient (Ric). To compare GOM and PINN performance, Ric coefficients were computed on 1,000 bootstrap resamples of the same dataset for each method. Statistical differences between methods were evaluated by paired comparison of the bootstrap Ric distributions. Also, Pearson’s linear correlations were included to examine the relationship between PINN-derived indices and the outputs of the global optimization method, when assessing real patient’s data. All statistical analyses were performed in R (v. 4.3.0) and p-values < 0.05 were considered significant.

## Results

### DIA-PINN performance on Synthetic PV-loop data

This section compares DIA-PINN and global optimization performance on synthetic pressure–volume loop data generated by Monte Carlo simulations.

DIA-PINN recovered accurate estimates of the prescribed values of the constitutive model, *θ*, under both single-beat and simulated vena cava occlusion conditions and irrespective of parameter bounding limit initialization. Using single-beat data, the error (Bias [95% CI]) of the PINN algorithm for *τ* was - 0.02 [-0.79 to 0.76] ms, for *V*_0_ was -0.05 [-2.1 to 2.0] mL and for *S*_+_ was -0.04 [-3.2 to 3.1] mmHg. However, the estimation of *S*_−_ exhibited wider dispersion and heteroscedasticity with an error of 0.34 [-4.45 to 5314] mmHg (carried over from the calculation of *V*_*d*_ with error: -1.5 [-20.1 to 17.7] ml).

In contrast, when fitting dummy vena cava occlusion instead of single beat, agreement improved markedly across all parameters. Bias was essentially null for *τ* (0.00 [-0.06 to 0.06] ms), (-0.06 [-2.3 to 2.2] mmHg), and *V*_0_ (-0.01 [-0.15 to 0.14]ml). Similarly, *V*_*d*_ showed reduced variability compared with the single-beat setting, yielding to more accurate values of *S*_−_: 0.02 [-3.48 to 3.52] mmHg (**Figure 2, Table 1 & Online Supplemental Table**). In both cases (single-beat and VCO) DAI-PINN overperformed GOM in recovering constitutive parameters as quantified by the infraclass coefficient across the Monte-Carlo-generated pool of synthetic LVs (**Figure 2, Table 1 & Online Supplemental Table**). Only when the parameter initialization and bounds were narrowly constrained a priori did the GOM achieve results comparable to the PINN (**Table 1**). *V*_*d*_, and therefore, *S*_*−*_, was shown to be dependent on the training data for both GOM and PINN approaches, yielding good performance only when *V*_0_ was larger than the minimum volume to fit (Bias [95% CI]: 0.01 [-0.34 to 0.35], see **Panel E in Figure 2 & Table 1)**.

**FIGURE 2:**
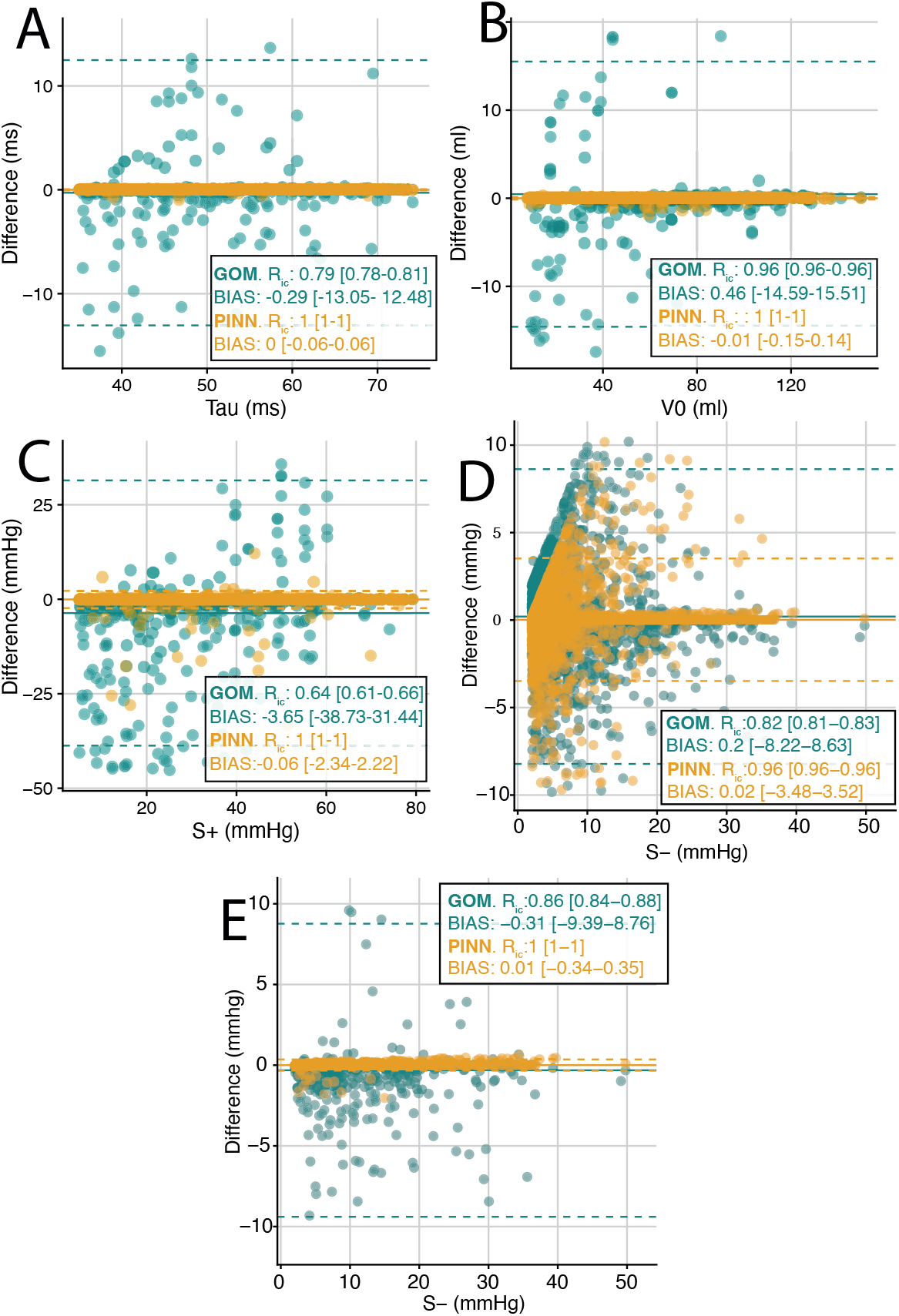
PINN performance to obtain diastolic properties from Monte Carlo simulations of vena cava occlusion using extended bounds for each method. Panels A to D, Bland-Altman plots for *τ*, *S*_+_, *V*_0_ and *S*_−_ respectively. Panel E, Bland-Altman plot for *S*_−_ using only PV data with *V*_0_ above end-systolic volume (ESV).

**Table 1.** Bootstrap computation of intraclass correlation coefficient (*Ric [95% CI]*) of the PINN and the GOM method to recover the value of each parameter of the Montecarlo Simulation for 1) single beat data, 2) vena cava occlusion using physiological bounds and 3) vena cava occlusion using extended bounds.

| | | $\tau$ | $V_0$ | $S_+$ | $S_-$ | $S_-$<br>($V_0 > ESV$) |
| --- | --- | --- | --- | --- | --- | --- |
| Single Beat | GOM | 0.97<br>[0.96-0.97] | 0.98<br>[0.98-0.98] | 0.97<br>[0.96-0.97] | 0.87<br>[0.86-0.88] | 0.93<br>[0.92-0.94] |
|  | PINN | 1<br>[1-1]* | 1<br>[1-1]* | 1<br>[1-1]* | 0.93<br>[0.92-0.93]* | 1<br>[1-1]* |
| VCO<br>Extended<br>bounds | GOM | 0.80<br>[0.76-0.80] | 0.96<br>[0.96-0.96] | 0.64<br>[0.62-0.64] | 0.82<br>[0.81-0.83] | 0.86<br>[0.84-0.88] |
|  | PINN | 1<br>[1-1]* | 1<br>[1-1]* | 1<br>[1-1]* | 0.96<br>[0.96-0.96]* | 1<br>[1-1]* |
| VCO<br>Physiological<br>Bounds | GOM | 0.99<br>[0.99-0.99] | 0.99<br>[0.99-0.99] | 0.98<br>[0.98-0.98] | 0.92<br>[0.92-0.93] | 0.95<br>[0.95-0.96] |
|  | PINN | 1<br>[1-1]* | 1<br>[1-1]* | 1<br>[1-1]* | 0.96<br>[0.96-0.96]* | 1<br>[1-1]* |

VCO: Vena cava occlusion. GOM: Global Optimization Method. PINN: Physics Informed Neural Network. ESV: End-Systolic Volume. \*: $p < 0.001$ on the paired comparison of the bootstrap $Ric$ distributions. VCO: Vena cava occlusion; $Ric$ : intraclass correlation coefficient; $\tau$ : time constant of left ventricular (LV) relaxation; $V_0$ : equilibrium volume; $S_+$ : constant of stiffness; $S_-$ : constant of elastic recoil.

In the single-beat approach, DAI-PINN converged after an average of 51,954 iterations (range: 14,381–89,527), requiring 1,573.32 seconds per run (range: 352–2,794). In contrast, for the vena cava occlusion, convergence was achieved in 1,400 iterations on average (range: 246–2,154), but with a substantially higher computational time of 48,628 seconds per run (range: 9,389–87,867).

DAI-PINN showed good performance for different parameter initializations. The mean absolute error (MAE) across repetitions was minimal for most parameters: 0.004 ms for *τ*, 0.064 mmHg for *S*_+_, 0.039 ml for *V*_0_, and 1 mmHg for *S*_*−*_ (ICC [95%CI]= 1[1-1], 0.99 [0.99-0.99], 0.99 [0.99-0.99] and 0.95 [0.94-0.95] respectively). These results indicate that the model consistently converged to similar parameter estimates regardless of the initial conditions, with only *V*_*d*_ showing relatively larger variability compared with the other parameters.

### Performance of the DAI-PINN on catheter PV loop measurements

DAI-PINN yielded similar results than the properly parametrized GOM when applied to clinical data. The relaxation time constant *τ* obtained by both methods were highly correlated (R=0.98, p<0.001, R_ic_: 0.98 [0.97-0.99]), and similar agreement was found for *V*_0_(R=0.99, p<0.001, R_ic_: 0.99 [0.98-0.99]) or *S*_+_ (R=0.90, p<0.001, R_ic_: 0.88 [0.83-0.92]). However, the elastic recoil coefficient *S*_*−*_ showed moderate correlation (R=0.77, p<0.001, R_ic_: 0.94 [0.91-0.96]), **Figure 3**), in line with our observations on synthetic data.

**FIGURE 3:**
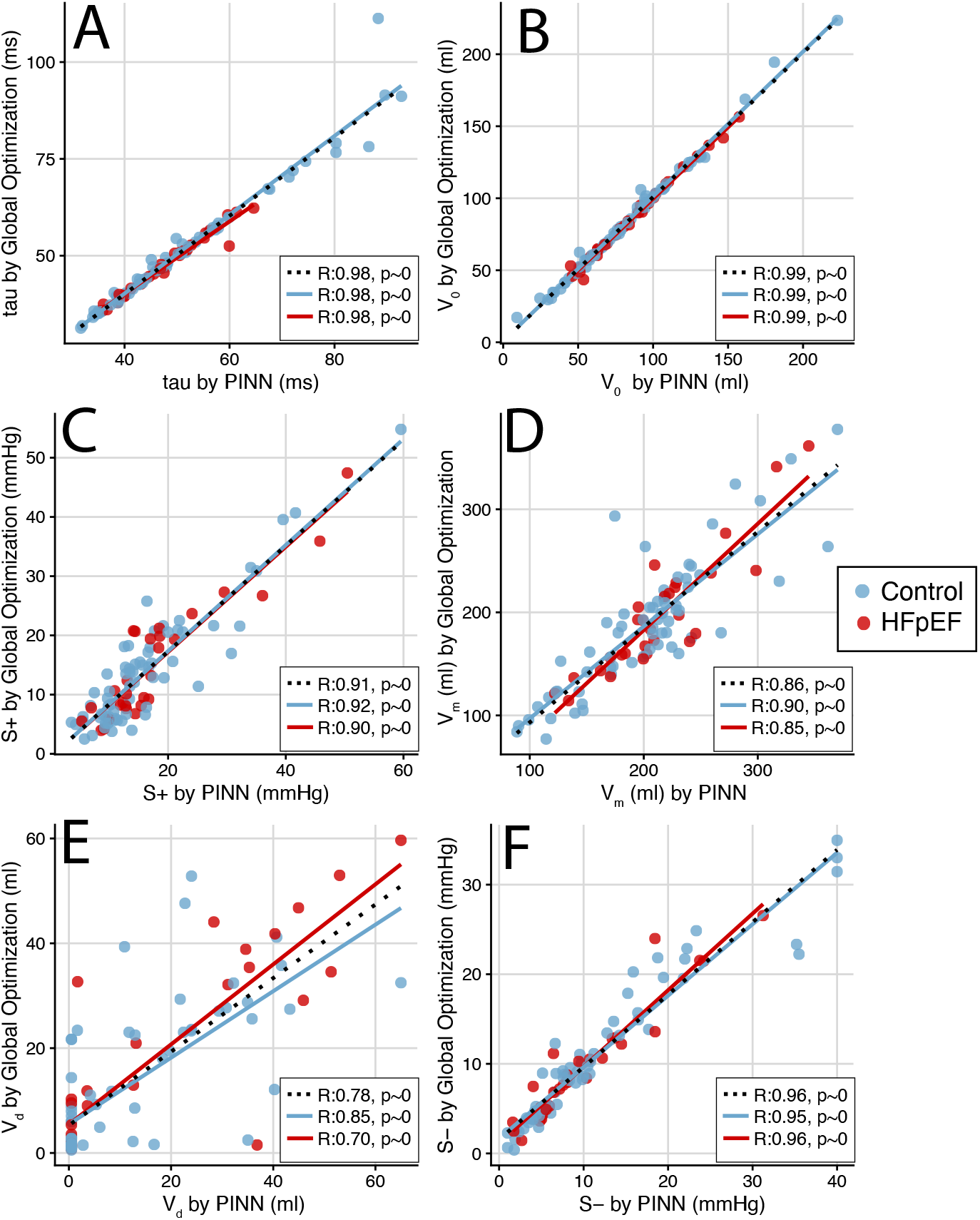
Relationship between the intrinsic LV properties in real patients assessed via global optimization method and the PINN. Panels A to E, plots for *τ*, *S*_+_, *V*_0_, *V*_*m*_, *V*_*d*_ and *S*_−_respectively. In red data from patients with HFpEF and in blue, data for the control group. Black lines represent fitting for the pooled groups.

Both methods also performed similarly for healthy controls and patients with HFpEF, with only small differences in the calculation of *S*_−_ (R=0.85, p<0.001 and R_ic_: 0.94 [0.89-0.97] for controls vs R=0.70, p<0.001, R_ic_: 0.94 [0.90-0.96] for patients with HFpEF).

## Discussion

The present study introduces DAI-PINN, a framework for estimating intrinsic diastolic properties of the left ventricle from pressure–volume data. Compared with prior approaches, the proposed method preserves the interpretability of constitutive modeling while improving numerical robustness and implementation versatility through a physics-informed learning framework, enabling simultaneous estimation of active relaxation, passive stiffness, and restoring forces across a wide parameter space.

### Performance to fit PV-loop data

Despite its shallow nature, proven enough to capture the main features of the model (28), the proposed PINN framework demonstrated high accuracy and robustness in recovering the constitutive parameters governing diastolic behavior, both in synthetic and clinical datasets. Across Monte Carlo simulations, the network achieved nearly unbiased parameter estimates and narrow limits of agreement, with intraclass correlation coefficients approaching unity for the main diastolic indices, improving GOM performance. When applied to clinical recordings, PINN-derived estimates were in excellent agreement with those obtained from the previously validated global optimization approach (14), yielding correlation coefficients and intraclass coefficients above 0.9 for the time constant of relaxation, *τ*, equilibrium volume, *V*_0_, and stiffness coefficient, *S*_+_. This performance confirms that the PINN can infer physiologically meaningful parameters directly from measured PV loops, maintaining consistency across different hemodynamic conditions and patient populations, including controls and HFpEF subjects.

Moreover, the framework is not restricted to the specific constitutive formulation adopted in this study. Any alternative diastolic or ventricular model—ranging from simpler exponential laws to more elaborate viscoelastic or time-varying elastance formulations, or those including the resting tone (11, 25, 29, 30) —can be readily embedded in the same PINN structure, offering a flexible and generalizable platform for modeling cardiac mechanics.

One key advantage of DAI-PINN approach over the GOM was its markedly reduced sensitivity to parameter initialization. While the GOM requires careful tuning of initial guesses and parameter bounds to ensure convergence and avoid spurious local minima (6), the PINN consistently converged toward the same parameter subspace across multiple random initializations. In fact, the mean absolute error across repetitions remained minimal for all parameters, with negligible dispersion in a wide range of estimates. This stability arises because the PINN integrates both data fidelity and physical constraints in its loss function, guiding convergence toward physiologically consistent solutions even from random starting points. Consequently, the method provides a more robust and reproducible framework for inferring diastolic properties, particularly when applied to noisy or incomplete datasets or under uncertain parameter initialization, where the GOM may fail or converge to non-physiological solutions.

As expected, the performance of DAI-PINN improved substantially when trained with full VCO sequences compared with isolated single-beat recordings. Under VCO conditions, the network is exposed to a wider range of loading states and dynamic pressure–volume relationships, which enhance parameter identifiability and reduce uncertainty in stiffness and equilibrium volume estimation. In contrast, single-beat inputs contain limited diastolic information and may lead to apparent overfitting, as the model attempts to reproduce the waveform with insufficient variability in volume and pressure data. This finding parallels classical load manipulation principles in PV analysis, where transient preload reduction provides the necessary mechanical diversity to decouple active and passive contributions. Therefore, while the single-beat PINN retains good accuracy, incorporating even modest beat-to-beat variability—as achieved during VCO—substantially improves the robustness and physiological interpretability of the inferred parameters.

### PINN-based cardiac modeling

Over the past few years, PINNs have gained popularity for their ability to integrate mechanistic models (e.g. partial differential equations or constitutive laws) into deep learning. In cardiac mechanics and hemodynamics, several studies have used PINNs to infer mechanical or electrophysiological parameters (31), often combined with patient-specific image-derived models (32). Some of these approaches include mechanical equations in weak form to estimate myocardial stiffness and active tension (33). Some works have used lumped models or reduced-order hemodynamic circuits embedded within a PINN to estimate parameters or predict flow rates. For instance, PINNs have been used in encoding a closed-loop circulation system embedding a left ventricle to rapidly estimate contractility parameters (17) or to map physiological time series (e.g. cuffless blood pressure) with minimal ground truth data, though in non-invasive peripheral settings (34). Despite their promise, these applications seldom—if ever—address the direct fitting of measured PV loop traces during diastole from patients and typically remain one step removed from raw clinical PV data. In contrast, the present work explicitly operates on real PV loop recordings, bridging the gap between theoretical PINN modeling and clinical ventricular functional assessment.

### Limitations

A key limitation of the present work, and ultimately of the constitutive equations used, concerns the identifiability of the dead-volume asymptote, *V*_*d*_, and subsequently of the elastic recoil index, *S*_−_, as defined by equation [3]. As shown in the Monte Carlo analyses, the PINN framework exhibited greater dispersion and heteroskedasticity in estimating *S*_*−*_, particularly when the fitted PV data corresponded to volumes above the equilibrium volume. This behavior arises because *V*_*d*_ primarily determines the passive pressure–volume relation in the low-volume asymptote. Thus, when the available data is far from this regime, the sensitivity of the pressure to *V*_*d*_ is minimal, and the corresponding gradients in the loss function are always zero. As a result, in this limit, *V*_*d*_ becomes mathematically non-identifiable, leading to multiple parameter combinations that yield nearly indistinguishable PV fitting and may affect the calculation of the elastic recoil constant. This phenomenon is well recognized in the broader field of cardiovascular modelling, where parameters that are not sufficiently “excited” by the data may become unidentifiable (35).

There are several strategies that may mitigate this limitation. First, data enrichment through preload reduction maneuvers, such as VCO, expands the sampled volume range toward and sometimes below *V*_0_, thereby steepening the sensitivity of the loss function to *V*_*d*_ and improving the calculation of *S*_−_. Also, it is important to incorporate physiological priors or inequality constraints as it may help to restrict the feasible parameter domain and avoid implausible solutions (36). Using models with alternative formulation for the passive pressure may be also useful to reduce parameter coupling.

## Conclusions

The proposed DAI-PINN accurately characterizes intrinsic diastolic properties of the left ventricle directly from measured pressure–volume loops, integrating mechanistic interpretability with data-driven robustness. This framework offers a flexible, initialization-insensitive, and physiologically consistent alternative to traditional methods, allowing broader clinical translation of physics-informed machine learning in cardiac mechanics.

## Funding Information

This work was partially supported by the Spanish Research Agency (AEI, grant number PID2023-146861OB-I00 to PML), Instituto de Salud Carlos III, Spain (grant number PI21/00274-PACER1 to JB) and the EU—European Regional Development Fund.

## Disclosures

All authors: Nothing to disclose.

## Data availability

Data supporting this study are available upon request. The source code used in this study is openly available at https://github.com/jftopham/PVLoop_PINN.

## Author Contribution

PML, JCdA and JB conceived and designed research; JFT and MGH performed experiments; JFT, MGH and PML analyzed data; PML and JB interpreted results of experiments; JFT and PML prepared figures; JFT and PML drafted manuscript; PML, JCdA and JB edited and revised manuscript, PML, JCdA and JB approved final version of manuscript.

## Notes

### Competing Interest Statement

The authors have declared no competing interest.

### Author Declarations

All procedures were conducted in accordance with the Declaration of Helsinki, and the research protocol was approved by the institutional ethics committee of Charite University Medicine Berlin (protocol number 225-07). Written informed consent was obtained from all participants prior to inclusion.

