## Supplementary Data for "DIA-PINN. A physics-informed machine learning method to estimate global intrinsic diastolic chamber properties of the left ventricle from pressure-volume data"

Javier Bermejo^1^,

Pablo Martinez-Legazpi^3^

^1^Department of Cardiology, Hospital General Universitario Gregorio Marañón; Facultad de Medicina, Universidad Complutense de Madrid, Instituto de Investigación Sanitaria Gregorio Marañón and CIBERCV, Madrid, Spain.

^2^Department of Mechanical Engineering and Division of Cardiology, Center for Cardiovascular Biology, University of Washington, Seattle, WA, USA.

^3^Department of Mathematical Physics and Fluids, Facultad de Ciencias, Universidad Nacional de Educación a Distancia, UNED and CIBERCV, Madrid, Spain.

***Brief Tittle***: Assessing LV Diastolic Properties using PINNs

**ONLINE SUPPLEMENTAL MATERIAL**

### ONLINE SUPPLEMENTAL FIGURES

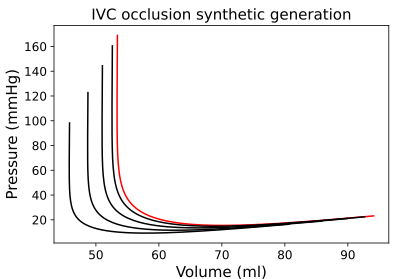

**Supplemental Figure SF1**: Synthetic pressure-Volume curves of a vena cava occlusion. Red line: Original PV data. Black lines vena cava occlusion progression.

### ONLINE SUPPLEMENTAL TABLE

**Online Supplemental Table ST1:** Bootstrap computation of intraclass correlation coefficient (Ric) of the PINN and the GOM method to recover the value of each parameter for single beat data and vena cava occlusion using physiological and extended bounds for each parameter.

|  |  | $V_{m}$ | $V_{d}$ | $V_{d}$  $(V_{0}>ESV)$ |
| --- | --- | --- | --- | --- |
| Single Beat | GOM | 0.96  [0.96-0.97] | 0.49  [0.48-0.49] | 0.90  [0.89-0.90] |
|  | PINN | 1  [0.99-1]* | 0.66  [0.64-0.66]* | 1  [1-1]* |
| VCO  Extended bounds | GOM | 0.49  [0.46-0.49] | 0.45  [0.42-0.48] | 0.67  [0.62-0.67] |
|  | PINN | 0.99  [0.99-0.99]* | 0.81  [0.80-0.81]* | 1  [1-1]* |
| VCO Physiological Bounds | GOM | 0.97  [0.96-0.97] | 0.66  [0.65-0.66] | 0.95  [0.95-0.95] |
|  | PINN | 0.99  [0.99-0.99]* | 0.81  [0.80-0.81]* | 1  [1-1]* |

VCO: Vena cava occlusion. GOM: Global Optimization Method. PINN: Physics Informed Neural Network. ESV: End-Systolic Voliume. *: p<0.001 on the paired comparison of the bootstrap Ric distributions.
